# Prevalence, intensity and risk factors of soil-transmitted helminthiasis after five effective rounds of preventive chemotherapy across three implementation units in Ondo State, Nigeria

**DOI:** 10.1101/2024.09.13.24313604

**Authors:** Hammed O. Mogaji, Francisca O. Olamiju, Fajana Oyinlola, Ijeoma Achu, Nimota O. Adekunle, Lydia E. Udofia, Ekaette G. Edelduok, Clement A. Yaro, Olanike O. Oladipupo, Alice Y. Kehinde, Fatai Oyediran, Moses Aderogba, Louise K. Makau-Barasa, Uwem F. Ekpo

**Affiliations:** Mission To Safe the Helpless, Lagos, Nigeria; Public Health Unit, Department of Applied Social Sciences, Marian University, Indianapolis, Indiana, United States; Department of Zoology and Environmental Biology, Olabisi Onabanjo University Ago-Iwoye, Ogun State; Department of Zoology, Akwa Ibom State University, Ikot Akpaden, Akwa Ibom State, Nigeria; Department of Animal and Environmental Biology, University of Uyo, Akwa Ibom State, Nigeria; Neglected Tropical Diseases Program Unit, Department of Public Health, Ondo State Ministry of Health, Akure, Ondo State, Nigeria; Neglected Tropical Diseases Program Unit, Department of Public Health, Federal Ministry of Health, Zonal Office, Akure, Nigeria; Neglected Tropical Diseases Program Unit, Department of Public Health, Federal Ministry of Health, Abuja, Nigeria; The Ending Neglected Diseases (END) Fund, New York, United States; Department of Pure and Applied Zoology, Federal University of Agriculture Abeokuta, Nigeria

**Keywords:** Soil-Transmitted Helminthiasis, Impact, Preventive Chemotherapy, Nigeria

## Abstract

**Background:** Routine epidemiological data are essential for monitoring the effectiveness of preventive chemotherapy (PC), optimizing resource allocation, and addressing the evolving needs in the elimination of soil-transmitted helminthiasis (STH). This study assesses the prevalence, intensity, and associated risk factors of STH following five rounds of albendazole-based PC in three implementation units (IUs) in Ondo State, Nigeria.

**Methodology:** Fresh stool samples were collected from 2,093 children aged 5–14 years across 45 systematically selected schools in three IUs: Ese-Odo, Irele, and Ile-Oluji. The samples were analyzed using the Kato-Katz technique. Additionally, standardized questionnaires were administered to gather data on demographics and access to water, sanitation, and hygiene (WASH) resources. Data analysis was conducted using R software version 4.3.2, with a 95% confidence interval.

**Principal Findings/Conclusions:** The parasitological data indicated a significant decline in the aggregated prevalence of STH across the three IUs. In Ese-Odo, the prevalence decreased to 25.8% (95% CI: 23.0–29.0) from 39% at baseline (d = -34%, p = 0.00). In Irele, prevalence dropped to 9.7% (95% CI: 7.6–12.0) from 51.3% at baseline (d = -81%, p = 0.00), and in Ile-Oluji, prevalence was reduced to 6.4% (95% CI: 4.6–8.7) from 23% at baseline (d = -72.2%, p = 0.00). The most prevalent STH species was *Ascaris lumbricoides*, with infection rates of 25.5%, 9.4%, and 6.4% in Ese-Odo, Irele, and Ile-Oluji, respectively, followed by *Trichuris trichiura* in Ese-Odo (2.7%) and Irele (0.4%), while hookworm infections were detected only in Irele (0.7%). The majority of infections were of low intensity in Ese-Odo (91.0%), Irele (96.8%), and Ile-Oluji (100%). Access to improved sanitation (17.7%, 54.9%, and 58.2%, p < 0.05), improved water sources (24.5%, 66.1%, and 69.8%, p < 0.05), and handwashing facilities (9.0%, 39.6%, and 25.4%) was suboptimal across Ese-Odo, Irele, and Ile-Oluji, respectively. Open defecation rates were high in Ese-Odo (54.2%), Irele (36.3%), and Ile-Oluji (34.3%). In Ese-Odo, significant risk factors for STH infection included the use of hand-pump boreholes (AOR: 2.44, 95% CI: 1.23–4.88, p = 0.01), unprotected dug wells (AOR: 3.25, 95% CI: 0.96–11.36, p = 0.06), ventilated improved pit latrines (AOR: 3.95, 95% CI: 1.13–16.1, p = 0.04), pit latrines without a slab (AOR: 2.19, 95% CI: 1.27–3.8, p = 0.01), and failure to use soap after defecation, both when soap was available (AOR: 12.09, 95% CI: 1.86–112.97, p = 0.01) and when soap was unavailable (AOR: 8.19, 95% CI: 1.73–76.65, p = 0.04). In Irele, access to protected dug wells was marginally significant (AOR: 1.79, 95% CI: 0.96–3.21, p = 0.06), while in Ile-Oluji, access to river water emerged as a significant risk factor (AOR: 7.97, 95% CI: 1.81–58.58, p = 0.02). The use of rainwater was found to be protective across all three IUs. These findings demonstrate significant progress in reducing STH prevalence across the three IUs following PC interventions. However, the data underscore the need for enhanced efforts to improve access to and use of WASH facilities to achieve STH elimination.

**Authors Summary:** The World Health Organization (WHO) recommends reducing soil-transmitted helminthiasis (STH) prevalence to below 20% and moderate-to-heavy infections to less than 2% in treated areas to achieve elimination targets. Endemic countries, including Nigeria, have implemented mass albendazole administration to at-risk populations to meet these goals. After a decade of such interventions in Nigeria, reassessing the infection status is essential for determining whether to adjust or discontinue the program. This study analyzed stool samples from 2,093 children across three implementation units (IUs) in Ondo State. The results indicated significant reductions in STH prevalence to below 20% in two of the three IUs, with no heavy infections reported in any IU. While these findings demonstrate the effectiveness of the PC interventions, the study also identified poor access to water, sanitation, and hygiene (WASH) resources, highlighting the need for improvements in these areas to sustain progress and prevent infection resurgence.

## Introduction

Soil-transmitted helminthiasis (STH) is a widespread yet preventable neglected tropical disease, affecting over 1.5 billion people globally, with a significant burden in sub-Saharan Africa [1-3]. STH is caused by four parasitic nematodes: *Ascaris lumbricoides* (roundworm), *Trichuris trichiura* (whipworm), *Necator americanus* (hookworm), and *Ancylostoma duodenale* (hookworm), which require soil for the development of their eggs [3,4]. Infection occurs primarily through the fecal-oral route, with individuals ingesting infective eggs of roundworm and whipworm or through skin penetration by hookworm larvae [3,4,5,6]. Transmission is often facilitated by inadequate infrastructure, particularly poor sanitation and hygiene practices, leading to contamination of water, soil, and food [7,8]. Rural and marginalized urban populations are particularly vulnerable, where sanitation and hygiene facilities are often absent or underutilized [9,10]. Young children, due to their developing immunity and increased exposure, face a higher risk of infection, resulting in adverse health outcomes such as malnutrition, anemia, impaired cognitive development, and educational disruptions [3,11-13]. These detrimental effects underscore the importance of preventive chemotherapy (PC), which involves mass administration of albendazole or mebendazole to at-risk populations [14].

The World Health Organization (WHO) has adopted PC as a primary strategy to control and eliminate STH [3,15]. Over the past decade, the WHO, in collaboration with two pharmaceutical companies GlaxoSmithKline and Johnson & Johnson, has coordinated the distribution of over 600 million donated medicines [16]. PC programs are implemented based on the prevalence of STH, determined through baseline epidemiological surveys in which biannual PC is recommended when prevalence exceeds 50%, annual PC is advised when prevalence is between 20–49.9%, and clinical case management is suggested when prevalence is below 20% [2,3,15,16]. It is anticipated that treating approximately 75% of the at-risk population over a period of 5–6 years will lead to a substantial reduction in STH endemicity [15,16]. Therefore, epidemiological surveys after five of effective (≥75%) rounds of PC are essential to evaluate program impact, optimize resource allocation, adjust or discontinue PC frequency, and mitigate the risk of developing drug resistance [2,3,15,16].

Nigeria, with its large population, is the most endemic country for STH compared to other sub-Saharan African countries [1,17-19]. Over the past eight years, the country has benefited from PC programs supported by donated medicines, and some areas are now presumed to have significantly reduced STH prevalence [20]. Given these developments, it is critical to conduct impact assessments to determine the current status of STH to decisions on future PC interventions. In this study, we evaluated the prevalence and intensity of STH following five effective rounds of albendazole PC interventions in three implementation units (IUs), corresponding to Local Government Areas (LGAs), in Ondo State, Nigeria.

## Materials and Methods

### Ethics

This study received ethical approval from the Ondo State Health Research Ethics Committee (approval number: NHREC/18/08/2016) and the Neglected Tropical Diseases Unit of the Federal Ministry of Health (reference number: NSCP/6/7/IV/23). Prior to data collection, we engaged key stakeholders from the Ministries of Health and Education at both the State and LGA levels in the study areas. This included organizing community meetings with representatives from the participating communities and schools. During these meetings, we provided detailed information to community leaders, teachers, parents, and children about the study’s objectives, procedures, and target population. Participation in the study was entirely voluntary and required informed consent from both the minors (aged 5–14 years) and their legal guardians. Children who agreed to participate provided written consent in the form of signatures or, where applicable, thumbprints on pre-printed informed consent forms (ICFs) on the day of sample collection. Only those children whose parents had previously provided consent, and who also received consent from a parent or teacher on the day of sample collection, were included in the study.

### Study area and settings

This study was conducted in the Ese-Odo, Irele, and Ile-Oluji Local Government Areas (LGAs) of Ondo State, located in Southwestern Nigeria. Ondo State comprises 18 LGAs with a population exceeding three million people. Baseline epidemiological mapping for soil-transmitted helminthiasis (STH) was conducted in 2013, revealing that three LGAs had endemicity levels below 20%, 12 had endemicity between 20% and 50%, and two LGAs had endemicity above 50% [20] (Fig. 1). Since 2013, the Federal and State Ministries of Health, in collaboration with partners such as EndFUND and Mission to Save the Helpless (MITOSATH), have implemented more than nine rounds of preventive chemotherapy (PC) with albendazole [20]. To date, an average of five effective PC rounds with therapeutic coverage exceeding 75% has been achieved. The three LGAs included in this assessment have met the minimum requirement of five to six effective PC rounds, making them eligible for an impact assessment survey.

**Fig 1:**
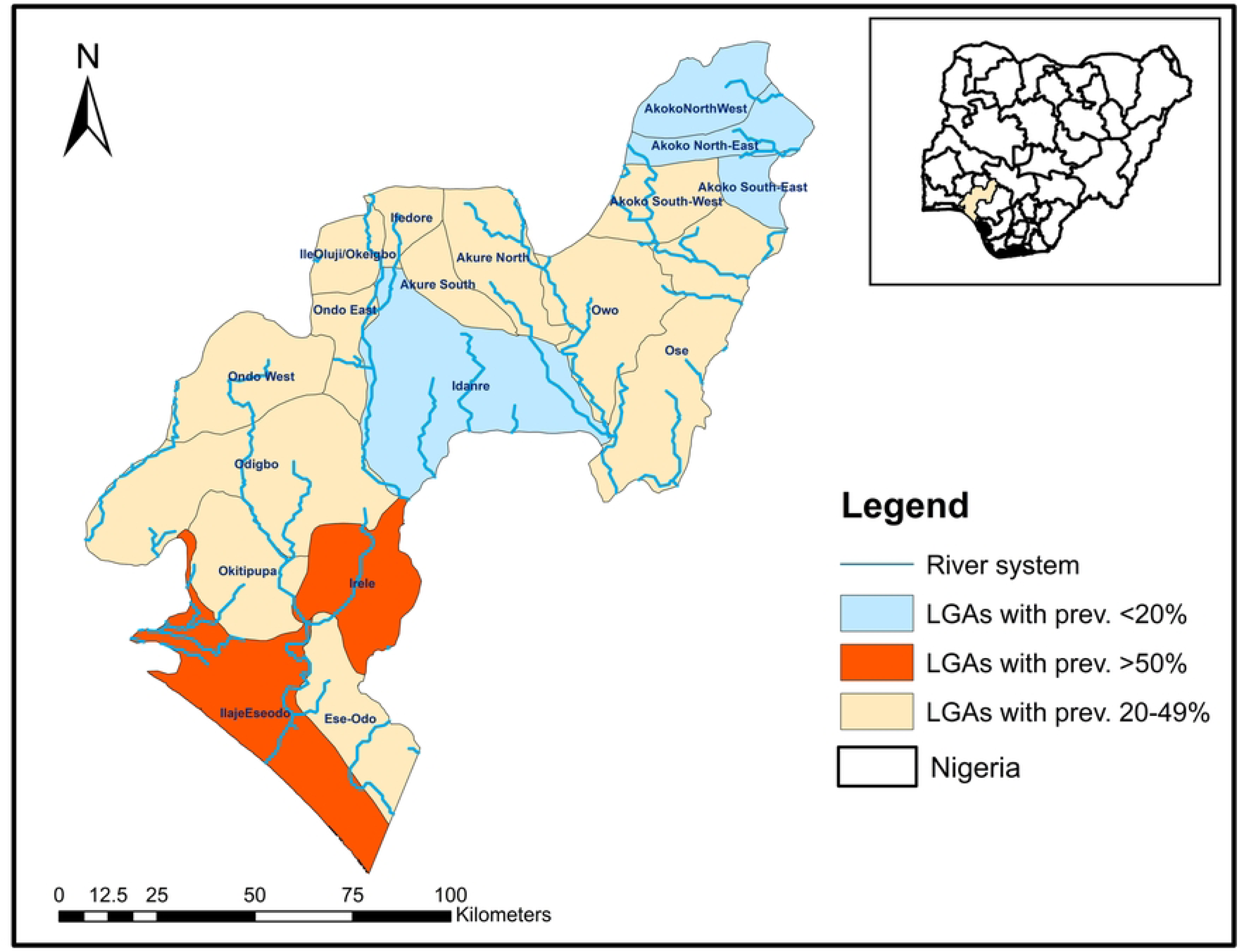
Map of Ondo State showing endemicity of the LGAs with Nigeria as inset.

### Study design and selection of sampling sites

This assessment utilized a cross-sectional sampling design conducted between March and April 2024. Data were collected through standardized questionnaires, and stool samples were obtained from school-aged children (SAC) across 45 systematically selected sampling sites (15 per LGA) in three Local Government Areas (LGAs). The sampling sites comprised government-owned primary schools, and participants (children aged 5–14 years) were selected following World Health Organization (WHO) guidelines [21]. Schools within each LGA were stratified by sub-districts (wards), and one school per ward was randomly selected using a paper ballot system from every five schools. This method ensured unbiased selection and adequate geographic representation across the LGAs and Wards [21].

### Sample size determination and selection of study participants

The World Health Organization (WHO) recommends recruiting 50 school-aged children (SAC) aged 5 to 14 years to assess worm burden and the impact of preventive chemotherapy (PC) [15]. However, recent guidelines suggest prioritizing 30 older SAC aged 10 to 14 years [21]. To comply with both sets of recommendations, we recruited a minimum of 50 SAC per school, including 30 late SAC (10–14 years) and 20 early SAC (5–9 years). A total sample size of 750 children was targeted for recruitment in each Local Government Area (LGA).

### Selection of study participants and exclusion criteria

Participants were recruited on school premises with the assistance of teachers. Children aged 5–9 years were selected from the first three primary school classes, with 6–7 boys and girls recruited per class. Children aged 10–14 years were selected from the fourth, fifth, and sixth primary school classes, with a target of at least 10 children (five boys and five girls) per class. A systematic selection process was employed: children were lined up by class and gender, assigned a number, and a sampling interval was calculated by dividing the total number of children by the required sample size per class. The first child was chosen through a paper ballot, and subsequent children were selected by adding the sampling interval to the previous child’s position. If the target sample size for a class was not met, additional children were recruited from the same class, school, or community. Exclusion criteria included children outside the target age range (5–14 years), those who were unwell, and those whose parents’ declined participation.

### Questionnaire administration

We utilized a closed-ended electronic questionnaire to collect epidemiological data from participants. Two types of questionnaires were administered via Android or iOS devices: (i) a school form and (ii) an individual questionnaire, both of which have been previously validated and employed in similar epidemiological surveys by our research team [10, 22, 23]. The school form captured information on school names, geographic coordinates, enrolment figures, and the availability of water, sanitation, and hygiene (WASH) facilities. The individual questionnaire collected demographic data, including age, gender, parental occupation, household WASH facilities, and parasitological results from the stool samples. Copies of the questionnaires are accessible at https://zenodo.org/records/11080490. All data were collected electronically and securely transferred to a remote backup server following each interview. Interviews were conducted in Yoruba and held confidentially, with a legal guardian or parent present when necessary.

### Collection of stool samples

Before administering the questionnaires, participants were required to complete informed consent forms and were assigned unique identification tags. These tags were affixed to their consent forms, sterile specimen containers, microscopic slides, and result sheets to maintain rigorous quality control throughout the data collection process. Participants were provided with applicator sticks, a plain sheet of paper, tissue paper, and soap to assist with stool collection. The voiding and collection process was supervised by a teacher and a member of the field team to ensure proper sample collection within the first hour of distribution and to prevent sample contamination or sharing. Bar soaps were distributed as incentives to promote good hygiene practices, with care taken to ensure that participants’ decisions to participate were voluntary and free from coercion.

### Parasitological assessment of stool samples

Stool samples were processed on-site within two hours of collection at a mobile laboratory set up by the field team. The Kato-Katz technique was used to process the specimens, and a single thick smear was prepared from each sample. After a clearing period of 30 minutes, the smears were examined microscopically for parasite eggs [24]. For quality assurance, each slide was re-examined by a second microscopist, and egg counts were verified [24]. A participant was classified as positive if at least one egg from any of the target parasites was detected per slide.

### Data Collection and Management

Data collection was remotely monitored using the KoboCollect platform, with field supervisors recording daily summaries through a closed-tracking tool developed in Microsoft Excel. This approach facilitated the triangulation of electronic uploads and enabled timely resolution of any discrepancies. Two distinct tracking tools were utilized during the study: one for school field forms and another for parasitology components. All collected data were imported into R Studio (version 4.3.2) for subsequent analysis. The data were encoded using a predefined codebook, with prevalence data recorded as numeric values—’0’ indicating no infection and ‘1’ indicating infection for each observed soil-transmitted helminth (STH) species with an egg count greater than one.

Overall infection status (i.e., any STH infection) was determined by summing values across rows using the ‘rowSums’ function. Infection intensity, expressed as eggs per gram (EPG) of stool, was calculated by multiplying the number of eggs observed per smear by 24. The intensity of *Ascaris lumbricoides* was classified as low (1–4,999 EPG), moderate (5,000–49,999 EPG), or high (≥50,000 EPG) [15]. For hookworms, intensities were classified as low (1–1,999 EPG), moderate (2,000–3,999 EPG), or high (≥4,000 EPG) [15]. *Trichuris trichiura* infections were categorised as low (1–999 EPG), moderate (1,000–9,999 EPG), or high (≥10,000 EPG) [15].

### Data analysis and interpretation

The analysis and interpretation of data for preventive chemotherapy (PC) adjustments followed the most recent World Health Organization (WHO) guidelines for STH prevalence impact assessment surveys [21]. Local Government Areas (LGAs) were categorised into four prevalence groups: (1) <2%, (2) 2% to <10%, (3) 10% to <20%, and (4) ≥20%. For LGAs in the first group, PC is recommended only during specific events, alongside the establishment and maintenance of surveillance systems. For LGAs in the second group, biennial PC over five years is recommended, with continuous surveillance. LGAs in the third group are advised to implement annual PC for five years with ongoing surveillance, while those in the fourth group are required to administer annual PC for five years. The aggregated proportion of moderate and heavy infections (MHI) was also calculated for each LGA, using a threshold of ≤2%, where the upper bound of the 95% confidence interval (CI) was below this limit.

The percentage change in prevalence between baseline and impact assessment surveys was also computed. To evaluate associations between variables, chi-square tests were employed, and Clopper-Pearson confidence intervals were calculated for prevalence estimates, where applicable. Univariate and multivariate logistic regression models were conducted to assess relationships between predictor variables (e.g., demographic characteristics and WASH conditions) and outcome variables (e.g., presence of any STH infection). Only variables with p≤0.05 in univariate analysis were included in multivariate models. Confounders and multicollinearity were examined using the Variance Inflation Factor (VIF), and variables with a VIF exceeding 5 were excluded. Crude and adjusted odds ratios (ORs) with 95% CIs were calculated, with a significance level set at P<0.05. Spatial distribution maps of STH across LGAs were generated using ArcGIS version 10.8. Further details on the dataset and analysis code can be accessed at https://zenodo.org/records/11080490.

## Results

### Profile of study locations and participants

Table 1 presents a comprehensive profile of the study locations across the three Local Government Areas (LGAs) of Ese-Odo, Irele, and Ile-Oluji. Each LGA consists of 10 sub-units, or wards, all of which were surveyed, achieving 100% geographic coverage. Sampling encompassed 22.1% of the 68 schools in Ese-Odo, 24.6% of the 61 schools in Irele, and 17.6% of the 85 schools in Ile-Oluji. Of the 750 school-aged children (SAC) targeted for recruitment in each LGA, 722 children (96.3%) were recruited in Ese-Odo, 750 children (100%) in Irele, and 621 children (82.8%) in Ile-Oluji. Stool samples were collected from 691 children in Ese-Odo (95.7%), 668 children in Irele (89.1%), and 613 children in Ile-Oluji (98.7%). The proportion of male participants exceeded that of females across all LGAs, comprising 54.0% in Ese-Odo, 50.4% in Irele, and 51.4% in Ile-Oluji. Additionally, a higher percentage of participants were in the 10–14-year age group: 54.8% in Ese-Odo, 59.6% in Irele, and 52.2% in Ile-Oluji (Table 1).

**Table 1:** Profile of study locations and participants.

### Prevalence and soil-transmitted helminthiasis across the LGAs

Table 2 provides a summary of the prevalence of soil-transmitted helminthiasis (STH) across the three surveyed Local Government Areas (LGAs): Ese-Odo, Irele, and Ile-Oluji. In Ese-Odo, 176 out of 691 individuals (25.5%) were infected with *Ascaris lumbricoides* (95% confidence interval [CI]: 22.0%–29.0%), with no cases of hookworm detected (0%). Nineteen individuals (2.7%) were found to have *Trichuris trichiura* infections (95% CI: 1.7%–4.3%). The overall prevalence of any STH in Ese-Odo was 25.8% (95% CI: 23.0%–29.0%). This represents a 34% reduction from the baseline prevalence of 39%, with a risk ratio of 0.66.

**Table 2:** Prevalence of soil-transmitted helminthiasis across the three LGAs and comparisons with the baseline survey.

In Irele, of the 668 individuals assessed, 63 (9.4%) were infected with *Ascaris lumbricoides* (95% CI: 7.4%–12.0%), five (0.7%) were found to have hookworm infections (95% CI: 0.28%–1.8%), and three (0.4%) were infected with *Trichuris trichiura* (95% CI: 0.12%–1.4%). The overall prevalence of any STH in Irele was 9.7% (95% CI: 7.6%–12.0%), indicating an 81% reduction from the baseline prevalence of 51.3%, with a risk ratio of 0.19.

In Ile-Oluji, out of 613 individuals surveyed, 39 (6.4%) were infected with *Ascaris lumbricoides* (95% CI: 4.6%–8.7%), with no cases of hookworm or *Trichuris trichiura* (0%, 95% CI: 0–0.78%). The overall STH prevalence in Ile-Oluji was 6.4% (95% CI: 4.6%–8.7%). This reflects a 72% decrease from the baseline prevalence of 23%, with a risk ratio of 0.28.

### Site-specific prevalence and spatial distribution of soil-transmitted helminthiasis across the three LGAs

Figures 2-4 display the endemicity maps for the three Local Government Areas (LGAs) included in the study. In 2011, Ese-Odo LGA was classified as endemic, with an overall prevalence of 39%, placing it in the 20–50% prevalence category. As a result, preventive chemotherapy (PC) was administered annually for five years. Although the current impact assessment showed a reduction in overall prevalence, the LGA remains within the same endemicity threshold (20–50%), with more than 50% of surveyed sites reporting a prevalence above 20% (Figure 2).

**Fig 2:**
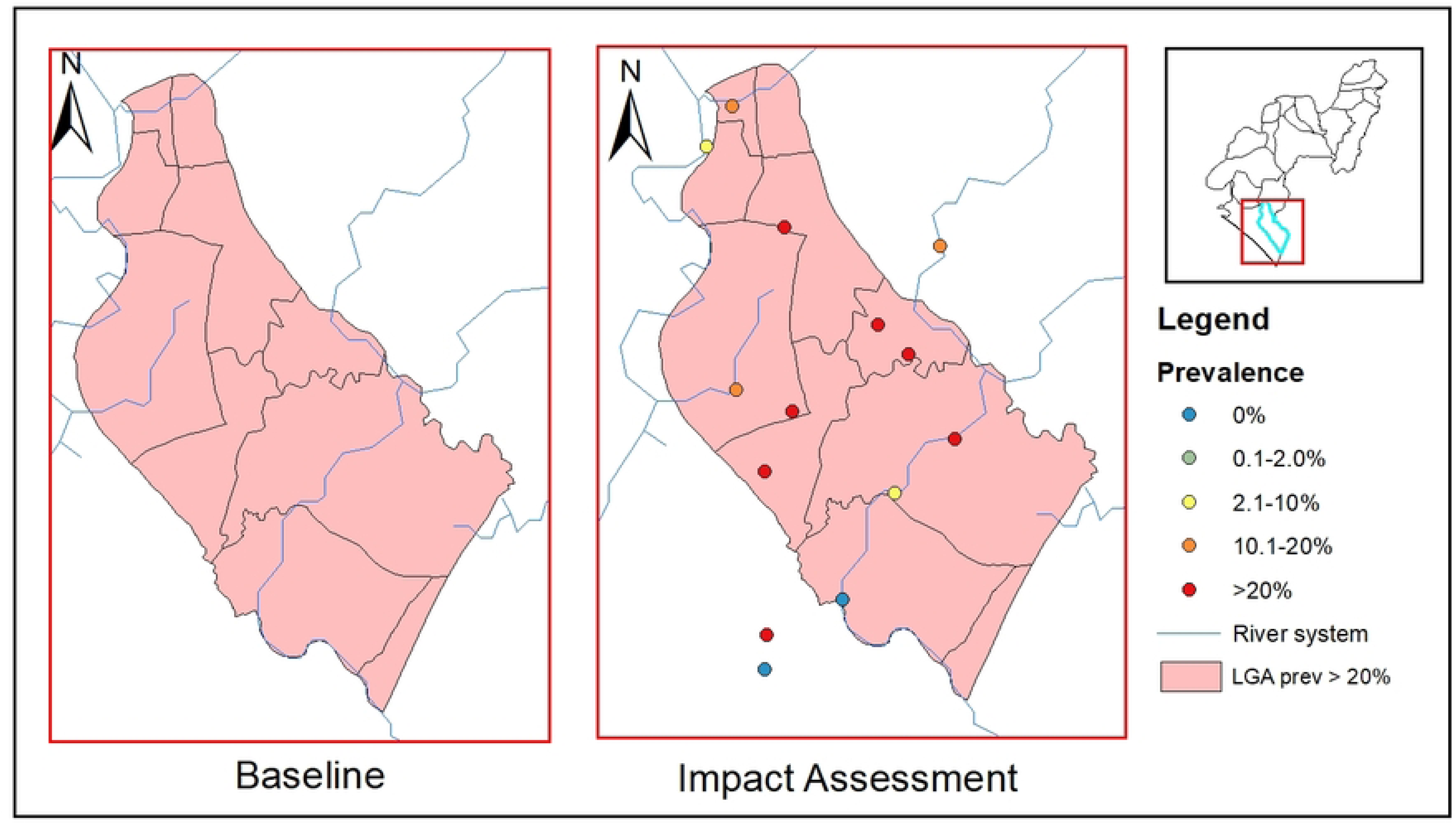
Endemicity map of soil-transmitted helminthiasis in Ese-Odo LGA at baseline and impact assessment survey.

**Fig 3:**
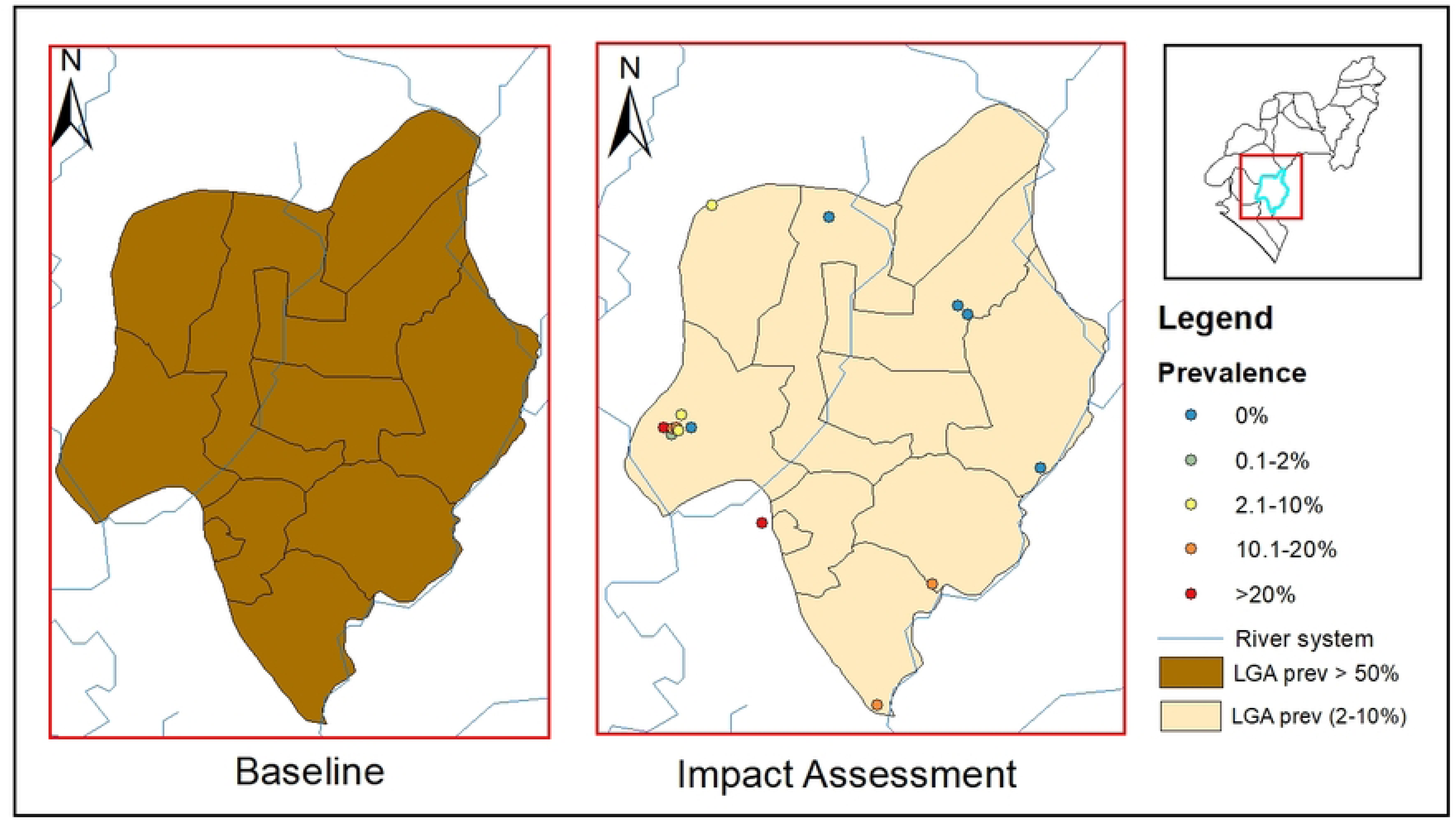
Endemicity map of soil-transmitted helminthiasis in Irele LGA at baseline and impact assessment survey.

**Fig 4:**
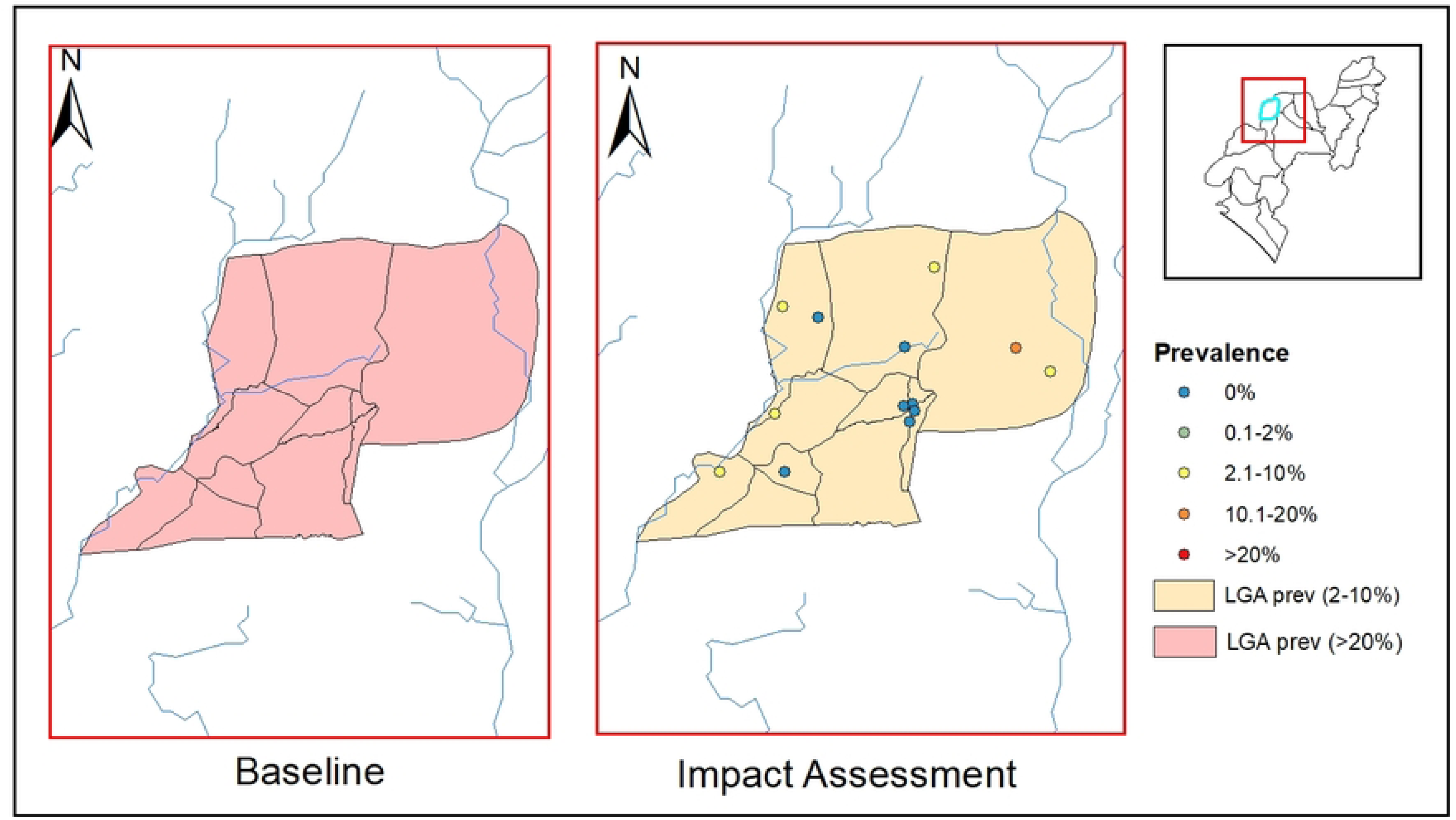
Endemicity map of soil-transmitted helminthiasis in Ile-Oluji LGA at baseline and impact assessment survey.

In Irele LGA, the baseline prevalence was 51.3%, placing it in the category requiring PC twice annually for five years. The current impact assessment revealed a significant reduction in overall prevalence to 9.7%, moving the LGA into the 2–10% prevalence category. Only 2 of the 15 surveyed sites in Irele reported a prevalence above 20% (Figure 3).

In Ile-Oluji LGA, the baseline prevalence was 23%, placing it in the 20–50% endemicity category, which required annual PC for five years. The current impact assessment indicated a decrease in overall prevalence to 6.4%, positioning the LGA within the 2–10% threshold. Similar to Irele, only 2 of the 15 surveyed sites in Ile-Oluji had a prevalence above 20% (Figure 4). Detailed site-specific prevalence data are provided in the supplementary file (S1).

### Intensity soil-transmitted helminthiasis infections across the three LGAs

Table 3 presents the distribution of potential morbidities associated with soil-transmitted helminth (STH) infections among the study participants. In Ese-Odo, 160 individuals (23.2%) infected with *Ascaris lumbricoides* had light infections, 15 (2.2%) had moderate infections, and one participant (0.1%) had a heavy infection. The 95% confidence intervals (CIs) for these infection levels ranged from 20% to 27% for light infections, 1.3% to 3.6% for moderate infections, and 0.01% to 0.93% for heavy infections (p=0.001). For *Trichuris trichiura*, 19 participants (2.7%) exhibited light infections, with no moderate or heavy infections observed (p=0.001).

**Table 3:** Intensity soil-transmitted helminthiasis infections across the three LGAs.

In Irele, 61 individuals (9.1%) infected with *Ascaris lumbricoides* had light infections, and two participants (0.3%) had moderate infections (p=0.001). The 95% CIs for light infections were 7.1% to 12%, and for moderate infections, 0.05% to 1.2%. *Trichuris trichiura* prevalence was very low, with only three participants (0.4%) exhibiting light infections, and no cases of moderate or heavy infections were detected (p=0.064). Additionally, among the five participants (0.7%) infected with hookworms, all had light infections, with no moderate or heavy infections reported (p=0.001).

In Ile-Oluji, 39 participants (6.4%) infected with *Ascaris lumbricoides* had light infections, with 95% CIs ranging from 4.6% to 8.7% (p=0.001). No cases of moderate or heavy infections were observed for any of the helminths.

### Status of water, sanitation and hygiene resources across the three LGAs

Table 4 details the status of water, sanitation, and hygiene (WASH) resources in the three local government areas (LGAs) studied. In Ese-Odo, the majority of the population, 579 individuals (80.2%), relied primarily on rivers as their main water source. Only a small fraction of participants had access to improved water sources, including protected dug wells (13.2%) and handpump boreholes (10.5%), while public taps were scarcely available (0.8%). Open defecation was a common practice, with 391 participants (54.2%) reporting this behavior. Pit latrines were widely used, with 119 participants (16.5%) utilizing pits without slabs and 66 participants (9.1%) using pits with slabs. A minority had access to flush toilets (5.8%) or ventilated improved pit latrines (2.8%). Regarding hygiene practices, only 65 participants (9.0%) reported having handwashing facilities in their toilets, and even fewer, 50 participants (6.9%), had access to soap within these facilities.

**Table 4:** Status of water, sanitation and hygiene resources across the three LGAs.

Similarly, in Irele, rivers were the most common water source, used by 442 participants (58.9%). Rainwater collection was another prevalent practice, reported by 329 participants (43.9%). Unprotected dug wells were used by 184 participants (24.5%), while public taps and handpump boreholes were available to 163 (21.7%) and 212 (28.3%) participants, respectively. Open defecation remained widespread, practiced by 272 participants (36.3%). Flush toilets were more commonly used in this LGA, with 220 participants (29.3%) reporting their use, followed by pit toilets with slabs (22.7%) and without slabs (19.5%). Handwashing facilities were available to 297 participants (39.6%), while 251 participants (33.5%) had soap in their toilets. A high proportion, 211 participants (84.1%), reported using soap after defecation.

In Ile-Oluji, rainwater was the primary water source for 366 participants (58.9%), followed by rivers, which were used by 339 participants (54.6%). Protected dug wells were another important water source for 275 participants (44.3%), though public taps (18.7%) and hand pumps (6.8%) were less frequently used. Open defecation was reported by 213 participants (34.3%). Among sanitation facilities, pit latrines with slabs were used by 154 participants (24.8%), flush toilets by 147 participants (23.7%), pit toilets without slabs by 68 participants (11.0%), and ventilated improved pit latrines by 60 participants (9.7%). Handwashing facilities were present in the toilets of 158 participants (25.4%), and 150 participants (94.9%) reported using soap after defecation, indicating a high level of hygiene compliance in this LGA.

### Association between demography, WASH and STH infections

Tables 5-7 present the results of logistic regression analyses evaluating the associations between sex, age, and water, sanitation, and hygiene (WASH) variables with the risk of soil-transmitted helminth (STH) infection. In Ese-Odo (Table 5), the adjusted model identified several significant risk factors, including the use of handpump boreholes (adjusted odds ratio [AOR]: 2.44; 95% confidence interval [CI]: 1.23–4.88; p=0.01), reliance on unprotected dug wells (AOR: 3.25; 95% CI: 0.96–11.36; p=0.06), the use of ventilated improved pit latrines (AOR: 3.95; 95% CI: 1.13–16.1; p=0.04), and pit latrines without slabs (AOR: 2.19; 95% CI: 1.27–3.8; p=0.01). Not using soap after defecation, despite having access to it, was also a strong risk factor (AOR: 12.09; 95% CI: 1.86–112.97; p=0.01), as was the absence or non-use of soap after defecation (AOR: 8.19; 95% CI: 1.73–76.65; p=0.04). The use of rainwater was identified as a protective factor against infection (AOR: 0.39; 95% CI: 0.24–0.64; p<0.001).

**Table 5:** Association between demography, WASH and STH infections in Ese-Odo.

**Table 6:** Association between WASH variable and STH infections in Irele.

**Table 7:** Association between WASH variables and STH infection in Ile-Oluji.

In Irele (Table 6), the adjusted model revealed that access to protected dug wells was a marginally significant risk factor (AOR: 1.79; 95% CI: 0.96–3.21; p=0.06), while the use of pit latrines without slabs was associated with a lower risk of infection (AOR: 0.38; 95% CI: 0.13–0.87; p=0.04).

In Ile-Oluji (Table 7), access to rivers was a significant risk factor for STH infection (AOR: 7.97; 95% CI: 1.81–58.58; p=0.02), while access to rainwater was found to be protective (AOR: 0.47; 95% CI: 0.22–0.97; p=0.05).

## Discussion

This study represents the first official impact assessment of soil-transmitted helminthiasis (STH) in Ondo State, following a decade of preventive chemotherapy (PC) interventions using albendazole. The importance of this assessment is underscored by several factors: first, to evaluate the efficacy of PC in reducing worm burden; second, to avoid the misallocation of resources to areas where they are no longer necessary, thereby maximizing returns on investment in underserved regions; and finally, to generate evidence necessary for measuring progress and optimizing PC programs for maximal effectiveness [15].

Recent guidelines from the World Health Organization (WHO) emphasize that for an area to meet elimination targets, all implementation units must achieve four critical conditions: reducing overall prevalence to below 20%, reducing the prevalence of moderate and heavy infections to less than 2% (including upper confidence intervals), reaching women of reproductive age (WRA), and improving access to water, sanitation, and hygiene (WASH), with a particular focus on eliminating open defecation [21]. While most country programs, including that of Ondo State, have concentrated on meeting the first two conditions, less attention has been given to the equitable allocation of resources for WRA and investment in WASH. The findings of this assessment underscore the need to expand PC interventions by integrating comprehensive WASH strategies.

The assessment found significant reductions in STH infections across the three local government areas (LGAs) evaluated. In Irele and Ile-Oluji LGAs, prevalence decreased to below 10%, with Irele experiencing an 81% reduction and Ile-Oluji a 72% reduction. Consequently, participants in Irele are now at only 0.18 times the risk of STH compared to baseline, while participants in Ile-Oluji are at 0.29 times the baseline risk. These results align with WHO recommendations of conducting PC every two years over five years, followed by continuous surveillance [15]. However, in Ese-Odo, the reduction in prevalence was modest, decreasing from 39% at baseline to 25.8% at the time of assessment, with prevalence still exceeding 20%. The risk of STH in Ese-Odo (66%) remained considerably higher than in the other LGAs, indicating a need for annual PC over the next five years [15]. Similar observations of limited progress despite PC interventions have been documented in countries such as Angola [28] and Kenya [29], where failures in implementation or lack of access to WASH have constrained program effectiveness [28,30].

While prevalence is commonly used to assess endemicity, infection intensity—measured by the number of parasitic eggs per gram of feces—is a more critical indicator of infection burden. Intensity levels provide a clearer indication of morbidity, as reductions in infection intensity correspond to diminished health risks from STH infections [26]. In line with WHO recommendations, the goal is to reduce the proportion of individuals with moderate or heavy infections to below 2% [15]. Among the three LGAs, only Ese-Odo had 2.3% of participants with moderate or heavy infections, predominantly due to *Ascaris lumbricoides*. Despite conducting six effective PC rounds (coverage >75%) out of nine since 2013, the high transmission rate in Ese-Odo raises concerns about demographic factors, WASH access, and the quality of implementation over the past decade [28].

Spatial analysis revealed that in Ese-Odo, more than 60% of sampled sites had prevalence rates ranging from 26% to 81%, with these high-prevalence areas widely dispersed. Baseline epidemiological surveys, conducted over a decade ago, used limited sampling strategies (typically five sites) within smaller frames to represent entire districts. Such methodologies reduce the ability to detect hotspots, particularly in areas with sparse or heterogeneously distributed infections [26,27]. Thus, the minimal reduction in prevalence observed in Ese-Odo may result from an underestimation of endemicity at baseline, a factor that influenced the decision to implement annual PC. Similar findings have been reported elsewhere, where outbreaks occurred in areas previously considered low endemic based on inadequate baseline data [30]. Consequently, robust assessment methodologies should be prioritized during PC program formulation to ensure that sampling strategies are comprehensive and effective.

A disaggregated risk analysis was conducted to examine the relationship between infection data and WASH variables. Social-ecological systems, particularly those related to WASH, are critical to the transmission of STH [10]. The study hypothesized that the limited reduction in prevalence and higher proportion of moderate to heavy infections may be linked to inadequate WASH access. The data supported this hypothesis, revealing poor access to improved sanitation, water sources, and handwashing facilities across the LGAs. Ese-Odo stood out, with only 17.7% of households using improved sanitation facilities, 25% using improved water sources, and just 9% having access to handwashing facilities. Open defecation rates were alarmingly high at 54.2%, much worse than in the other LGAs. Regression models indicated that inadequate hygiene practices, such as failure to use soap after defecation, presented the highest risk for STH infection, followed by the use of unimproved latrines and water sources. These findings align with recent reports from Ethiopia [32] and highlight the need for targeted interventions to improve WASH access and hygiene practices within these communities.

From a policy perspective, these findings underscore the need for timely impact assessments after five effective PC rounds, as recommended by WHO, to prevent resource wastage. Routine continuation of PC without evaluating its effectiveness leads to unnecessary expenditures in drug procurement and program operations, diverting funds from other health priorities. While past arguments pointed to the lack of standardized methods for impact assessment as a challenge, the recent WHO Monitoring and Evaluation (M&E) framework for schistosomiasis and STH provides clear guidelines for evaluation [21]. Donors and partners should thus prioritize impact assessments to enhance programmatic decision-making, prevent resource misallocation, and optimize the value of their investments. Further research into the comparative costs of conducting impact assessments versus continuing routine PC without evaluation is needed to provide stronger evidence of cost-efficiency. The present data offer clear evidence of the financial savings that can be achieved through timely evaluations, reinforcing the importance of sustainable and impactful disease control programs.

## Conclusion

The implementation of the preventive chemotherapy (PC) program for soil-transmitted helminthiasis (STH) has proven highly effective in Irele and Ile-Oluji, with prevalence rates reduced to less than 10%. However, in Ese-Odo, the reduction in prevalence has been limited to 66%, with current rates still around 25% and moderate-to-heavy infections persisting at 2.3%. The continued transmission of STH in Ese-Odo a riverine LGA is largely attributed to inadequate access to and use of water, sanitation, and hygiene (WASH) facilities. Given these findings, a shift to biennial PC in Irele and Ile-Oluji is recommended, while annual PC should be maintained in Ese-Odo for the next five years. Additionally, targeted investments in WASH infrastructure, with a priority focus on Ese-Odo, are crucial to complement PC efforts and facilitate progress toward STH elimination as a public health issue. Finally, this study emphasizes the importance of conducting impact assessments after five effective rounds of PC, as recommended by WHO, to optimize resource allocation and ensure the sustainability of control programs.

## Data Availability

Date used in the study can be accessed via https://zenodo.org/records/11080490

## Acknowledgements

We extend our heartfelt gratitude to the community leaders, health workers, education secretaries, and staff of the NTDs Control Department, Ondo State Ministry of Health, for their unwavering support and cooperation throughout this study. We sincerely acknowledge the contributions of individuals who served as field supervisors, technicians, interviewers, and members of the technical support team. We also appreciate the broader health community for their support and collaboration in this endeavor. The efforts of the following field workers are duly acknowledged: Jacob Babasola Ajayi, Oluwaseun Bunmi Awosolu, Oyinkansola Suliat Fadiji, Azeez God’sgift Ibrahim, Adedotun Ayodeji Bayegun, Cynthia Uchechukwu Umunnakwe, Olajumoke Olubukola Taiwo, Olubukola Deborah Adelakun, Effiong Okon Odiongenyi, Victor Kayode Gboyega, Stephen Ayodeji Oluwole, Ifeoluwa Adeniyi George, Promise Funmilayo Akinwale, Uzodimma Ikechukwu Vincent, Ridwan Olamilekan Mogaji, Oladotun Joseph Bankole, Damilola Rachael Adeleye, Boluwatife Ifeoluwa Adebayo, Abdulrahman Afolamade Aladejana, Orosunnegan Dunsin Elizabeth and Uwem Aniefiok Essien. Their dedication and hard work were instrumental to the success of this impact assessment.

## Authors’ contributions

UFE, FOO, and HOM conceptualized the study; HOM prepared the protocol, whereas UFE, FOO, IU, and FOO improved the protocol; HOM, UFE, FOO, FO, NOA, LEU, EGE, CAY, and OOO participated in supervised field surveys and data collection; HOM performed all statistical analyses; HOM prepared the first draft of the manuscript; HOM, UFE, FOO, IU, FO, MA, and LM contributed to the development of the final manuscript and approved its submission.

## Availability of data and materials

The datasets used and analyzed during the current study have been attached as supplementary files [See here at https://zenodo.org/records/11080490.].

